# Fine-tuned large language models enhance influenza forecasting

**DOI:** 10.1101/2025.03.27.25324747

**Authors:** Chenxiang Li, Qiqiao Zhang, Yue Zhang, Bowen Zhao, Jule Yang, Li Qi, Jun Ding, Dechao Tian

## Abstract

Influenza-like illness (ILI) continues to present significant challenges to global health, highlighting the need for accurate forecasting to guide timely public health responses. Traditional statistical and deep learning models, though widely applied, often face difficulties in capturing complex nonlinear dynamics and addressing data scarcity. This study examines the potential of fine-tuned large language models (LLMs), including Llama2 and GPT2, for multi-step influenza forecasting. A specialized fine-tuning framework is introduced, incorporating custom embeddings and a prediction block, and evaluated on seven real-world ILI surveillance datasets. Extensive benchmarking against SARIMA, LSTM, and PatchTST demonstrates that fine-tuned LLMs consistently deliver superior accuracy and stability, with particular advantages in long-term forecasts. Pre-trained LLMs, while able to capture broad temporal patterns in zero-shot scenarios comparable with SARIMA, gain substantial improvements in precision through fine-tuning. These results establish fine-tuned LLMs as practical and robust solutions for influenza forecasting, offering new opportunities to enhance epidemic modeling in data-limited public health environments.

## Introduction

Influenza-like illness (ILI) remains a significant public health challenge, causing widespread illness and straining healthcare systems worldwide. Each year, seasonal influenza leads to an estimated one billion cases and 290,000 to 650,000 respiratory-related deaths globally [1]. Influenza forecasting, the prediction of future influenza trends based on historical data, supports public health decision-making by providing early warnings, guiding resource allocation, and enabling timely interventions to reduce the societal burden of influenza.

Influenza forecasting has been extensively studied. Traditional models, such as the seasonal autoregressive integrated moving average (SARIMA) [2], remain widely used due to their strong theoretical foundation and effectiveness in simple, linear time series settings. However, these models struggle with nonlinear, non-stationary patterns and perform poorly for long-term forecasts, limiting their utility for seasonal influenza planning. Machine learning and deep learning models, including long short-term memory (LSTM) [3] networks and Transformer-based models [4], have been increasingly adopted for capturing complex temporal dynamics and long-range dependencies. However, their reliance on large-scale datasets presents challenges for influenza forecasting, where datasets are often small-scale and typically consist of weekly summary reports. This mismatch highlights the need for models that can operate effectively on limited data while accurately modeling complex epidemic dynamics.

The rapid evolution of large language models (LLMs) has transformed natural language processing and now extends to complex tasks beyond textual analysis. Recent studies demonstrate the emerging potential of LLMs in public health research, including detecting RNA viruses [5], predicting inpatient and intensive care unit mortality [6], leveraging multilingual social media analytics for emergency public health decision-making [7], and enhancing health care communication [8]. However, their application in influenza forecasting remains limited, underscoring the need for systematic exploration and rigorous validation.

This study explores the fine-tuning of pre-trained, open-source LLMs, including Llama2 and GPT2, for forecasting influenza activity. The task is formulated as a multi-step time series forecasting problem, where historical data from the preceding 52 weeks are used to predict influenza activity over the following 13 weeks. To exploit the sequential learning capabilities of LLMs, originally developed for token prediction in natural language processing, we developed a specialized fine-tuning architecture. This architecture incorporates modifications specifically designed for influenza forecasting. It includes a customized embedding layer that transforms the temporal input sequence into a dense matrix representation, capturing sequential patterns, and a fully connected prediction module that outputs future influenza activity values (Figure 1).

**Figure 1:**
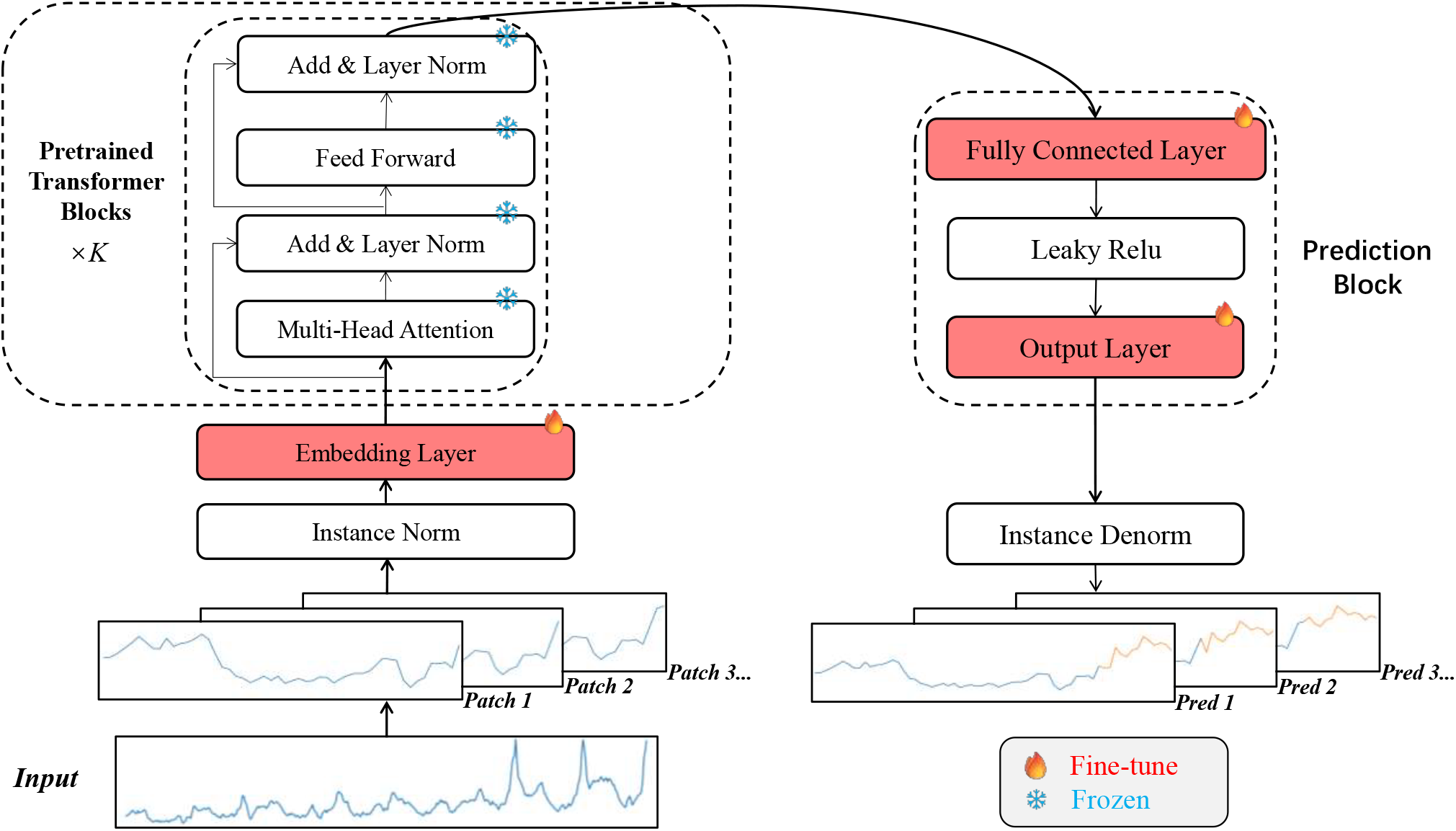
Fine-tuning architecture for LLMs. The input time series data is divided into overlapping patches for independent training and evaluation. Each patch undergoes instance normalization and embedding layer before passing through frozen pre-trained transformer blocks composed of multi-head attention and feed-forward layers. A task-specific prediction block then converts the learned representations of each patch into predictions. Fine-tuning is limited to the embedding layer, fully connected feed-forward layer, and output layer (highlighted in red), enabling task adaptation while preserving the pre-trained LLM’s general knowledge.

The fine-tuning architecture is applied to seven real-world influenza surveillance datasets spanning 2010 to 2020 (Supplementary Figure S1). Each dataset is partitioned into training, validation, and testing subsets. A one-step sliding window approach generates overlapping patches in the training data, allowing each time point to appear in multiple shifting contexts while preserving temporal order. This helps the model learn both short-term fluctuations and long-range dependencies. During fine-tuning, parameters in the pre-trained LLMs are frozen; only those in the embedding layer and prediction module are updated (Figure 1). Model performance is evaluated on the testing subset and benchmarked against widely used statistical and deep learning baselines, including SARIMA, LSTM, and the recently developed PatchTST model (Methods).

Using the ILI positive rate for the 13-week window before the COVID-19 pandemic in Chongqing, China as a case study, fine-tuned Llama2 predictions closely align with observed values, achieving the lowest mean absolute error (MAE) (Figure 2). Notably, Llama2 accurately predicts the first 10 weeks, except for the second week, while other models struggle with the fourth and ninth weeks. In this case study, fine-tuned Llama2 consistently demonstrates superior prediction accuracy across multiple error metrics, including mean squared error (MSE), mean absolute percentage error (MAPE), symmetric mean absolute percentage error (SMAPE), and trend agreement measured by Spearman’s correlation coefficient. Fine-tuned GPT2 exhibits the highest trend agreement as quantified by Pearson correlation coefficient (Table S1). Together, these findings in the case-study showcase the advantages of fine-tuned Llama2 and GPT2 in predictive accuracy compared to alternative methods.

**Figure 2:**
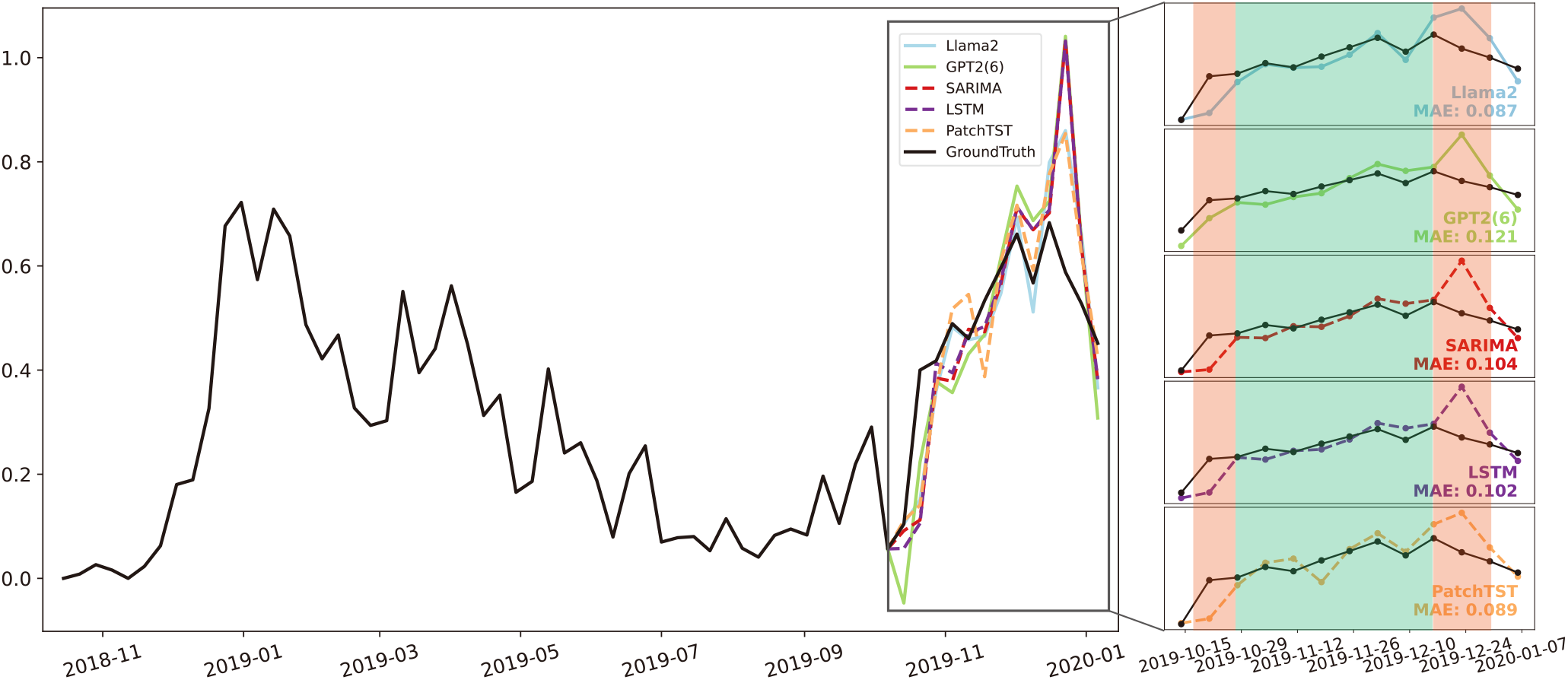
Case study of predicting ILI positive rates in Chongqing, China from October 2019 to January 2020. A 52-week observation window spanning November 2018 to September 2019 is used to predict ILI positive rates for the subsequent 13 weeks. The inset plot zooms in on predicted versus observed values, with large deviations highlighted in red and close alignments in green. Among five modelsfine-tuned Llama2, fine-tuned GPT2, SARIMA, LSTM, and PatchTSTLlama2 shows the lowest MAE and best performance.

Building on the initial case study, a systematic evaluation is conducted through a sliding-window forecasting approach, where a 52-week window is incrementally advanced by one week to predict the subsequent 13-week window in the testing datasets (Methods). Fine-tuned Llama2 and GPT2 consistently demonstrate superior performance compared to other models. Across the seven datasets, fine-tuned Llama2 and GPT2 achieve the lowest MAE in six out of seven cases, highlighting their predictive accuracy. Fine-tuned Llama2 achieves the lowest MAE in three ILI positive rate datasets: Northern China, Chongqing, China, and the USA. Fine-tuned GPT2 achieves the lowest MAE in ILI positive rate and ILI case datasets from Southern China, as well as ILI cases from Northern China. LSTM performs best in terms of MAE for the remaining ILI case dataset from Chongqing, China (Figure 3a). Further bench-marking using additional three error-based metrics, namely MSE, MAPE, and SMAPE, reinforces the superior performance of fine-tuned Llama2 and GPT2 across datasets. PatchTST and SARIMA achieve the best results in specific scenarios, additional to LSTM (Table S3, Table S4). Summarizing performance across the four error-based metrics and seven datasets shows that both fine-tuned Llama2 and GPT2 individually achieve the highest accuracy in 9 out of 28 cases, followed by LSTM and SARIMA with 4 cases each, and PatchTST with 2 cases (Figure 3b). Fine-tuned Llama2 and GPT2 not only compete with other models but also closely rival each other, prompting an evaluation that considers both the best and second-best performances. Once again, fine-tuned Llama2 and GPT2 demonstrate clear advantages over alternative models (Figure 3c). Togetehr, fine-tuned Llama2 and GPT2 consistently emerge as the most accurate models, highlighting the robustness of fine-tuned LLMs in influenza forecasting.

**Figure 3:**
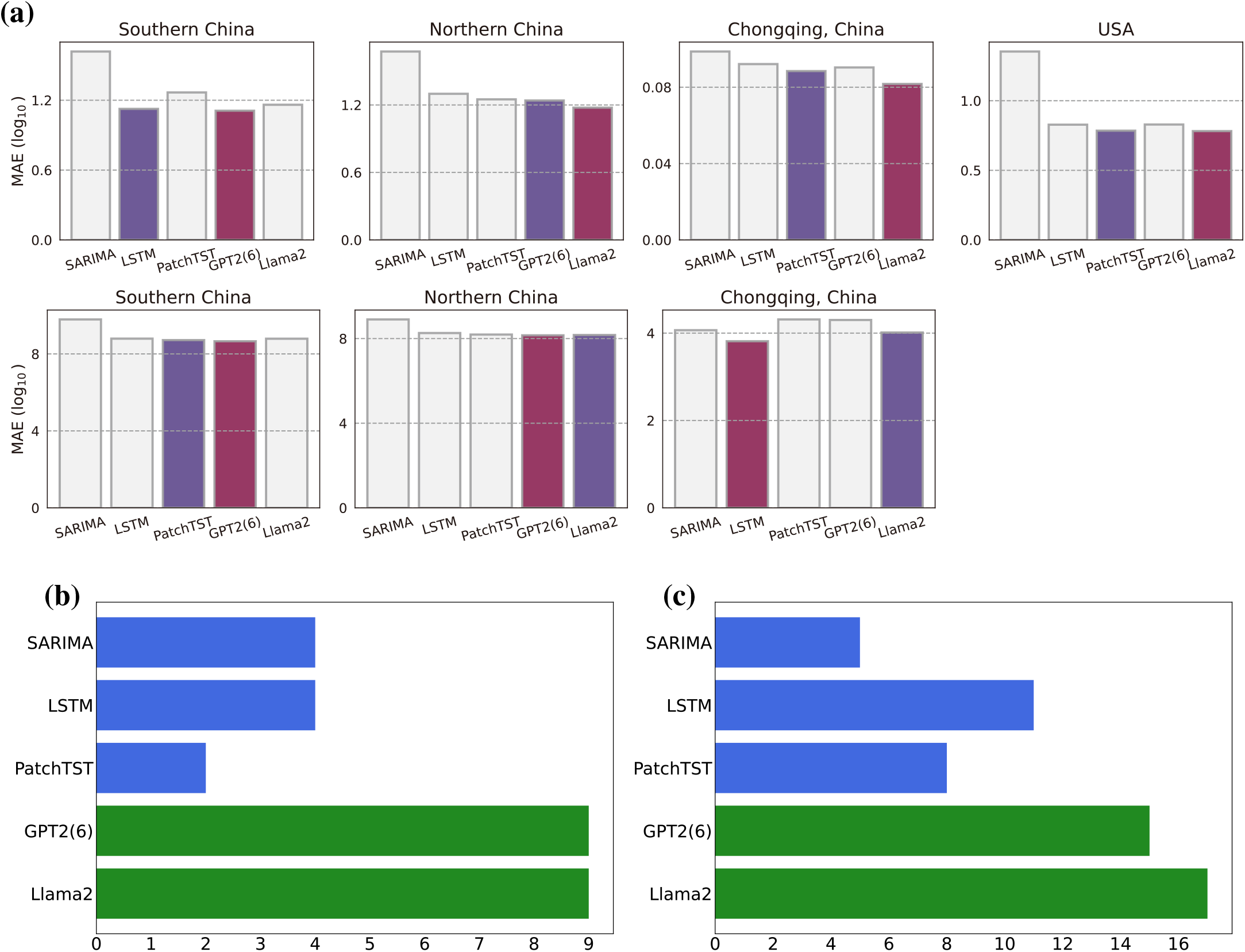
Benchmarking predictive performance across seven datasets. (**a**) MAE comparisons, with the best-performing bars highlighted in red and second-best in blue. The top row displays results for ILI positive rate prediction using four datasets, while the bottom row shows results for ILI case counts prediction using three datasets. (**b**) Number of top-ranked results obtained by each model across four error metrics (MAE, MSE, MAPE, and SMAPE) evaluated on seven datasets, yielding a total of 28 combinations. (**c**) Number of top-ranked and second-ranked results obtained by each model across all evaluations. Fine-tuned LLMs are shown in green, and alternative models in blue.

Different public health applications require varying prediction lengths. Short-term forecasts are essential for outbreak detection and immediate intervention, while longer-term predictions support resource planning and epidemiological trend analysis. Evaluating model performance across prediction lengths from 1 to 13 weeks provides insights into forecasting consistency and stability. Fine-tuned Llama2 exhibits the highest accuracy and stability, particularly for longer predictions (4 to 13 weeks). LSTM, GPT2, and PatchTST follow in descending order of predictive performance, each maintaining consistent accuracy across varying prediction lengths. In contrast, SARIMA exhibits significant instability, with drastic fluctuations across prediction lengths, though it performs best for next-week predictions (Figure 4). Instability may stem from the model’s linear assumptions, leading to accumulated errors and difficulties in capturing complex dependencies over long forecasts. LLMs, in particular, excel at capturing nonlinear patterns, ensuring consistent performance across different forecasting windows compared to traditional models. These results highlight the advantage of fine-tuned Llama2 in delivering stable long-term forecasts, reinforcing its role as a reliable tool for influenza activity forecasting.

**Figure 4:**
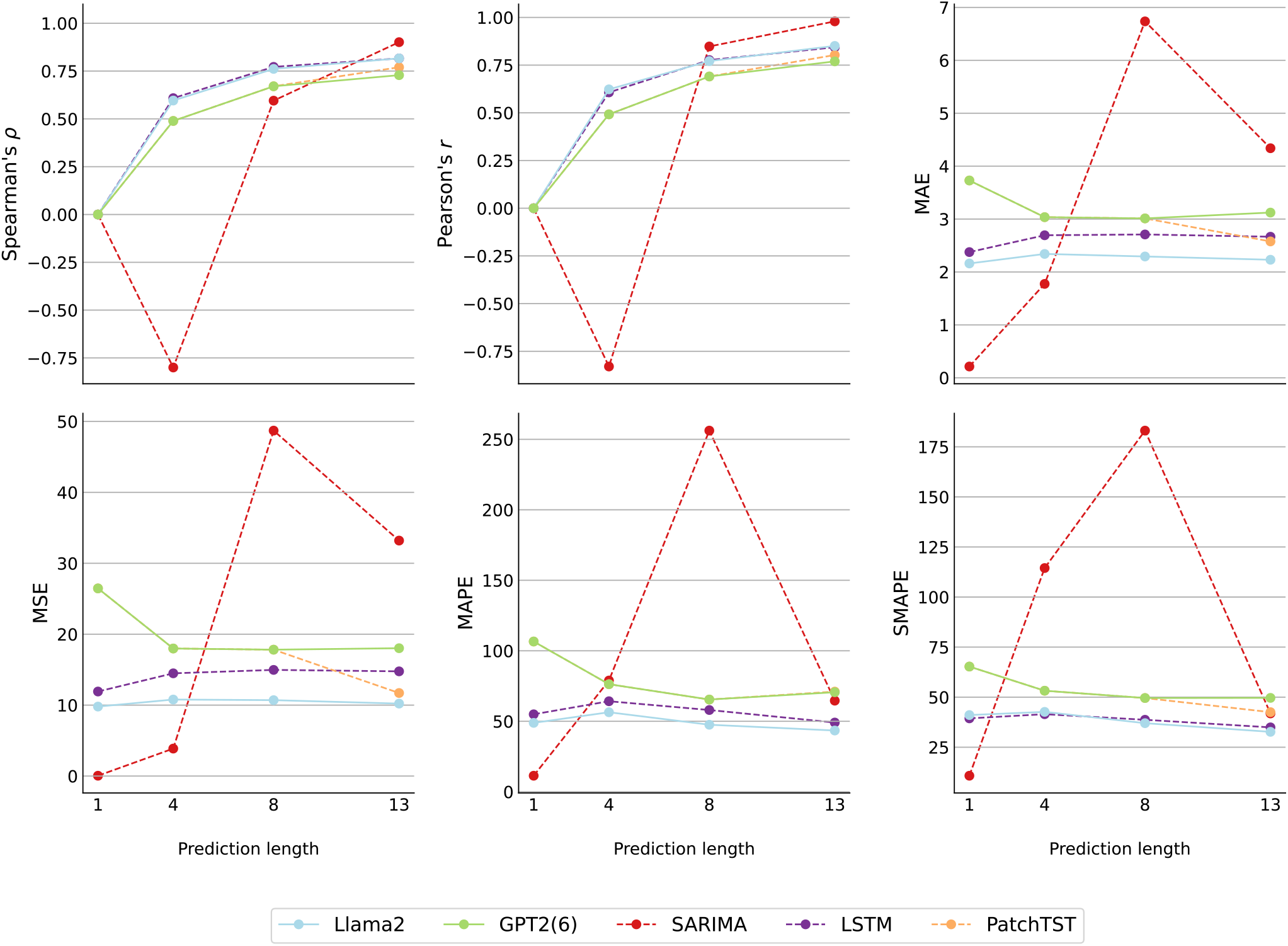
Benchmarking model performance across varying prediction lengths. Model performance is evaluated on the ILI positive rate dataset from Northern China over prediction lengths ranging from 1 to 13 weeks, using six metrics: Spearman correlation coefficient, Pearson correlation coefficient, MAE, MSE, MAPE, and SMAPE. Spearman and Pearson correlation coefficients are set to zero for a prediction length of one, where correlation is undefined.

LLMs have shown remarkable zero-shot capabilities across various domains, motivating their evaluation for influenza forecasting. To do this, following the same experimental framework established for benchmarking fine-tuned LLMs against alternative methods, pre-trained LLMs are directly applied to forecast 13-week future intervals based on 52-week historical observations in a sliding-window manner in the testing data. The only difference is that pre-trained models are used without fine-tuning on training data, ensuring a fair and consistent comparison. When compared to SARIMA across seven datasets, pre-trained Llama2 and GPT2 show comparable trend metrics (median change in Spearman and Pearson correlations: 1.74% and –0.05%) and only marginal increases in error (median change in MAE: 1.04%, MSE: 11.28%, MAPE: 2.96%, SMAPE: 2.48%; Supplementary Table S12 and S13). Against LSTM and PatchTST, pre-trained Llama2 and GPT2 show higher trend correlations (median Spearman and Pearson increases of 15.44% and 17.28% for Llama2; 13.87% and 17.88% for GPT2) but also higher errors (MAE: 86.22% and 109.95%; MSE: 218.98% and 251.38%; Supplementary Table S14, S15, S16, and S17). Compared to their fine-tuned counterparts, pre-trained Llama2 and GPT2 similarly achieve higher Spearman and Pearson correlations (7.61% and 14.71% for Llama2; 10.27% and 17.22% for GPT2), but incur substantially higher errors (MAE: 95.67% and 110.37%; MSE: 240.43% and 279.68%; MAPE: 55.72% and 51.28%; SMAPE: 32.58% and 12.85%; Supplementary Table S18 and S19). Collectively, these findings indicate that while pre-trained LLMs effectively capture broad influenza trends and perform similarly to SARIMA in both trend and error metrics, fine-tuning is essential for more accurate and reliable ILI forecasting.

Ablation experiments assess the impact of different model components by systematically altering them to evaluate their contribution to performance. The number of layers in pre-trained LLMs is a key factor. GPT2 performs best with the first six out of 12 layers, aligning with Zhou et al. [9], while Llama2 achieves optimal results using all 32 layers (Supplementary Result A.2). Further analysis shows that training fully connected (FC) layers in the prediction block enhances forecasting accuracy, while retraining layer normalization and positional embeddings offers minimal improvement. Reducing model weight precision to 8-bit maintains performance, making it viable for resource-limited settings. A learning rate of 1 *×* 10^−4^ is optimal (Supplementary Result A.2). However, larger models such as Llama3, Gemma2, and prompted-fine-tuned GPT4-o mini did not demonstrate clear improvements and often underperformed when fine-tuned under the same architecture (Supplementary Tables S2, S3, S4). This suggests that the limited scope of influenza surveillance data (approximately 523 weekly observations from 2010 to 2020) restricts the effective utilization of larger model parameters.

To evaluate the feasibility of fine-tuning LLMs in resource-limited environments, execution time and memory consumption were assessed across multiple models (Table 1). Experiments were conducted on a Tesla T4 GPU (16 GB) using the Northern China influenza dataset. Fine-tuning GPT2 (6-layer) for 64 epochs required approximately 3.6 minutes, while Llama2 took about 24.8 minutes. Memory usage remained within practical limits, with GPT2 peaking at 3.08 GB of CPU memory and 0.58 GB of GPU memory, and Llama2 at 2.93 GB of CPU memory and 12.47 GB of GPU memory. The higher resource demand for Llama2 reflects its substantially larger parameter size. Fine-tuning newer models such as Llama3 and Gemma2 is also feasible under the same fine-tuning architecture. Although computational costs are higher than those of traditional deep learning models, they remain acceptable for practical applications, reinforcing the feasibility of fine-tuning LLMs in real-world scenarios.

**Table 1:** Training parameters and computational cost comparison on positive rate dataset from northern China. The experiments are conducted using a Tesla T4 GPU with 64 training epochs.

| Model | # Tuning Parameters | # Frozen Parameters | Time (min) | Peak CPU Mem (GB) | Peak GPU Mem (GB) |
| --- | --- | --- | --- | --- | --- |
| Llama2 | 39.9M | 5B | 24.75 | 2.93 | 12.47 |
| Llama3 | 39.9M | 6.98B | 23.77 | 3.43 | 12.55 |
| GPT2 | 7.5M | 85M | 3.62 | 3.08 | 0.58 |
| Gemma2 | 34.9M | 8.32B | 94.77 | 3.13 | 12.20 |
| PatchTST | 20M | 0 | 4.07 | 3.11 | 0.59 |
| LSTM | 21.5K | 0 | 0.19 | 3.45 | 0.02 |

In conclusion, compared to traditional models such as SARIMA, LSTM, and PatchTST, fine-tuned LLMs (e.g., Llama2 and GPT2) consistently show better forecasting accuracy across seven datasets, highlighting their ability to capture complex temporal patterns. These results indicate that fine-tuned LLMs are practical and reliable for influenza forecasting. In public health settings where large, curated datasets are often unavailable, pre-trained LLMs offer a clear advantage by drawing on knowledge from diverse training corpora. As model design, fine-tuning methods, and domain-specific foundation models continue to improve, fine-tuned LLMs have the potential to become essential tools in influenza forecasting and broader epidemiological modeling.

## Methods

### Pre-trained LLMs evaluated for influenza forecasting

Two widely used open-source pre-trained LLMs, GPT2 [10] and Llama2 [11], are extensively tested. In addition, more recent LLMs, including Llama3, Gemma2, and GPT-4o mini, are also evaluated to explore the potential for influenza forecasting. Pre-trained parameters for these models are downloaded from Hugging Face (Appendix A.1). The fine-tuning architecture of the LLMs, depicted in Figure 1, includes several tailored modifications to optimize their performance for influenza forecasting.

### Influenza data preprocessing

The input historical observational data, denoted as *y*_1_, *y*_2_, …, *y*_*T*_, represents the observed influenza data at each time point *t*. The data undergoes first-order differencing:

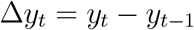

and 12-step seasonal differencing:

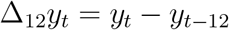

to remove trends and seasonality, which is a common practice for influenza forecasting.

The patching technique [12] is applied to the preprocessed data, which is divided into overlapping patches, enabling more comprehensive model training, particularly when the sample size is limited. To improve the model’s stability and speed up convergence, the input influenza series is normalized using its mean and variance:

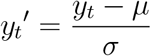

Here, *µ* is the mean and *σ* is the standard deviation of the influenza series.

For a fair comparison, the preprocessed data are used as the input for all models, including fine-tuned LLMs and alternative methods, ensuring consistency in evaluation and eliminating variability due to differences in data handling.

### LLMs fine-tuning architecture

Despite normalization, the time-series data still differs significantly from regular text data used for LLMs. To address this, the original embedding layer of the pre-trained LLMs is replaced with a fully connected layer, leveraging linear probing to reduce computational requirements. Given that the input *Y* is in the form of a tensor *Y* ∈ *ℝ*^*b×*52*×*1^, representing a batch of *b* samples with 52 time steps, the transformations are applied as follows:

First, the input *Y* undergoes a rearranging operation to transform its shape from ℝ^*b×*52*×*1^ to ℝ^*b×*1*×*52^. This rearrangement ensures that the data aligns with the expected input format for subsequent operations:

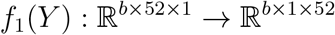

Second, padding is applied along the last dimension to make the sequence length compatible with the input requirements of the following layers:

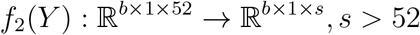

Here, *s* is the padded length.

Next, the padded tensor *Y* is unfolded into smaller local segments of length corresponding to the patch size:

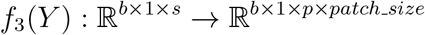

This operation slices the data into patches, with *p* denoting the number of patches and each patch having a size of *patch size*

Finally, the patches are rearranged to prepare for embedding layer:

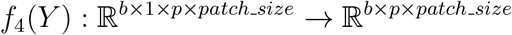

The embedding of the input data is then computed via a linear transformation:

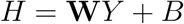

where *H* is the resulting embedding, **W** is the weight matrix, and *B* is the bias term.

According to Zhou [9], self-attention layers and feedforward neural networks (FFN) capture most of the knowledge from pre-trained LLMs. While their approach fine-tunes the embedding, normalization, and output layers while freezing the self-attention blocks, this leaves a large number of parameters, potentially leading to inefficiencies. To address this, all parameters of the pre-trained LLMs are frozen, simplifying fine-tuning and reducing computational costs.

In the prediction block, a fully connected layer with a Leaky ReLU activation function is applied before the output layer for influenza forecasting tasks. The compelte process is defined as:

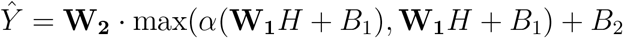

where 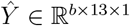 is the predicted output, representing the influenza forecasts for the next 13 weeks. *H* is the output from the LLM, which serves as the learned representation of the data. It captures important patterns such as trends, seasonal variations, and any other underlying temporal dependencies in the data. *W*_1_ and *W*_2_ are the weight matrices corresponding to the two layers of the fully connected network. *W*_1_ maps the input features to an intermediate space, and *W*_2_ further transforms this intermediate representation to the final output, which predicts the influenza rate. *B*_1_ and *B*_2_ are the corresponding bias terms. These biases allow the model to adjust the output independently of the input data, providing more flexibility in learning. Lastly, *α* is a small constant (typically *α* = 0.01) that controls the slope for negative values of *H*. The Leaky ReLU activation function ensures that the model does not completely discard negative values, allowing it to learn from negative activations while preventing issues like the vanishing gradient problem.

Final predictions are obtained by applying inverse normalization and inverse differencing to the models output. This architecture integrates the strengths of pre-trained LLMs with domain-specific adjustments, enabling efficient and accurate influenza forecasting.

### Experimental setup

For the influenza positive rate and ILI data, results for 13-step ahead predictions are presented using the GPT2-backbone Frozen Pretrained Transformer and fine-tuned Llama2-backbone models as examples. Due to space limitations, only these results are shown, while complete experimental results for all models can be found in Table S2, Table S3, and Table S4.

We use GPT2 to represent GPT2-backbone with first 6 hidden layers, and Llama2 to represent Llama2-backbone with all 32 hidden layers. The influenza series, such as ILI cases, are split into training, validation, and test sets in a 7:1:2 ratio. The forecasting process involves using data from the past 52 timesteps to predict the next 13 timesteps. The default loss function is computed using the mean squared error (MSE) as follows:

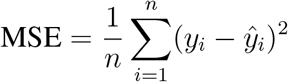

where *y*_*i*_ is the actual value, 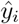 is the predicted value, and *n* is the number of samples. This procedure does not apply to the SARIMA model. For this classical statistical approach, the configuration described by Feng et al. [13] is followed, fitting the SARIMA model on all data prior to the final 13 weeks and making predictions for those final 13 weeks. Consequently, when evaluating models by calculating metrics, the results for other models (except SARIMA) represent the average performance on the test set (except Figure 2).

### Model deployment and privacy considerations

To ensure data privacy and address potential security concerns, we conducted all experiments using locally deployed LLMs. Specifically, we utilize open-source models GPT2 and Llama2, selected as representative architectures with hidden state dimensions of 768 and 4096, respectively. To accommodate differences in model architecture, we tailor the input projection layers to align the input embeddings with each model’s hidden dimensionality. These modifications are applied uniformly across all experimental settings to preserve implementation consistency and support rigorous comparative evaluation. For the proprietary GPT-4o mini model, we removed all time-related metadata from the input to prevent leakage of sensitive temporal information. However, due to the closed-source nature of GPT-4o mini, we were unable to perform detailed internal analysis as with open-source models. In response to this constraint, we designed specific prompts to guide its behavior and ensure controlled outputs. A standardized prompting format is used (see Appendix A), enabling consistent evaluation while maintaining data privacy.

### Benchmark analysis

#### Alternative methods

The performance of the proposed LLM-based models is assessed by comparing them with classical and state-of-the-art models commonly used in influenza forecasting. The comparison models include **SARIMA**[13], a robust and interpretable classical statistical model that remains widely applied. Since the official implementation is not available, we reimplemented it based on the methodology described by Feng et al.; **LSTM**[14], a recurrent neural network model designed for sequential data modeling and well-suited for capturing temporal dependencies, which is constructed using PyTorch’s *torch*.*nn* module; and **PatchTST**[12], a Transformer-based model that utilizes advanced self-attention mechanisms and demonstrates strong performance in time series forecasting tasks. We adopted the implementation provided by Zhou et al., available at https://github.com/DAMO-DI-ML/NeurIPS2023-One-Fits-All.

#### Evaluation metrics

The evaluation is performed on influenza positive rate and ILI datasets, with prediction lengths set to 8 and 13 steps ahead. Six evaluation metrics are employed for a comprehensive comparison: Spearman correlation coefficient (Spearman’s *ρ*), Pearson correlation coefficient (Pearson’s *r*), MSE, MAE, MAPE, and SMAPE. The formulas for all evaluation metrics, except for MSE which has been defined previously, are given below:

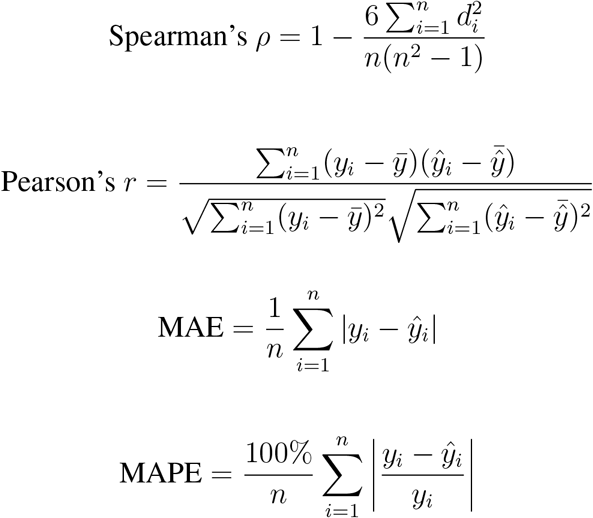

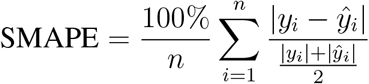

where *y*_*i*_ represents the actual values, 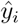 denotes the predicted values, 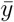 and 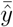 are the means of the actual and predicted values respectively, *d*_*i*_ is the difference between the ranks of *y*_*i*_ and 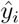, and *n* is the number of samples.

## Data availability

Among the seven ILI datasets used in this study, five influenza virological and ILI surveillance datasets are public data and obtained from Feng et al. [13], which were collected by the National Influenza Surveillance Network in China and the US Centers for Disease Control and Prevention (CDC). These datasets cover the period from April 2011 to March 2020 and reflect influenza activity trends and positive test rates, based on over 3.7 million samples from China and over 8.3 million samples from the US. Additionaly, two real-time weekly datasets, including influenza positive rates and ILI case counts from January 2010 to January 2020 in Chongqing, China, are provided by the Chongqing CDC. These datasets are not publicly available but can be accessed upon reasonable request.

## Code availability

The code is available on GitHub: https://github.com/licx11/LLMs4Influenza.

## Acknowledgement

This work was supported by the National Natural Science Foundation of China grant 12271536, Shenzhen Sustainable Research grant KCXFZ20211020172545006, and Shenzhen Science and Technology Program grant ZDSYS20230626091203007 and Guangdong Basic and Applied Basic Research Foundation grant 2022A1515010043 to D.T, partially supported by NSERC RGPIN2022-04399 to J.D. Additionally, this work was supported by High-performance Computing Public Platform (Shenzhen Campus) of Sun Yat-sen University.

