## Supplementary File for "Fine-tuned large language models enhance influenza forecasting"

### A Supplementary appendix

#### A.1 LLMs Applied in This Paper

The following pre-trained models, sourced from Hugging Face and Kaggle, are employed in this paper:

- **GPT-2**: https://huggingface.co/openai-community/gpt2
- **Llama2**: https://huggingface.co/meta-llama/Llama-2-7b-hf
- **Llama3**: https://huggingface.co/meta-llama/Meta-Llama-3.1-8B-Instruct
- **Gemma-2**: https://www.kaggle.com/models/google/gemma-2/transformers

While forecasting influenza series using GPT4-o mini, we follow the setup proposed by Gruver et al. [15], utilizing the following prompt template:

```
{"messages": [{"role": "system", "content": "You are a helpful
   assistant that performs time series predictions. The user will
   provide a sequence and you will predict the remaining sequence.
   The sequence is represented by decimal strings separated by commas
   ."}, {"role": "user", "content": "Please continue the following
   sequence without producing any additional text. Do not say
   anything like \u2019the next terms in the sequence are\u2019, just
    return the numbers. Sequence:290, -2897, 1073, 192, 362, -4,
   2285, 1158, 182, -1897, -2832, 909, -1499, 645, -204, 2071, 379,
   -626, -647, -1235, -350, -394, -5932, -1540, -3091, -3398, -2404,
   1745, 9731, 7249, 8215, 7744, -759, 2112, 542, -5590, 3542, 2822,
   6613, -4568, -6377, -5180, -3796, -3109, 275, -781, -802, -426,
   409, -1015, -354, 648"}, {"role": "assistant", "content": "-400,
   599, -330, 136, 592, 143, -1021, -1993, 825, 1315, 2087, -637,
   1650"}]}
{"messages": [{"role": "system", "content": "You are a helpful
   assistant that performs time series predictions. The user will
```

```
provide a sequence and you will predict the remaining sequence.
The sequence is represented by decimal strings separated by commas
."}, {"role": "user", "content": "Please continue the following
sequence without producing any additional text. Do not say
anything like \u2019the next terms in the sequence are\u2019, just
 return the numbers. Sequence:145, -74, 140, 19, 108, 16, 8, -20,
102, -135, -90, 61, 89, -165, -15, 46, 134, 65, 62, -71, -28, 20,
-56, 144, -33, -181, 102, -74, -77, -25, -49, 177, 82, -118, 57,
84, 12, -16, 244, -106, 100, 0, 16, -14, 10, -198, 16, 31, -178,
142, -237, -52"}]}
```

#### A.2 Supplementary Results

##### *Evaluation of Layer Configurations in GPT2 and Llama2*

In this section, we evaluate how the number of GPT2 and Llama2 layers affects performance, thereby providing justification for our choices of fine-tuned GPT2 and fine-tuned Llama2. According to Zhou et al.[9], the pre-trained GPT2 model achieves optimal results when using between three and nine layers, leading them to adopt the six-layer variant as their default. Building on this insight, we conducted experiments on the positive rate dataset from Northern China, comparing different GPT2 and Llama2 layer configurations (see Figure S2 and Figure S3). Our findings on GPT2 align with those reported by Zhou, confirming that the 6-layer configuration is indeed optimal. However, when evaluating Llama2, performance continued to improve as the number of hidden layers increased, prompting us to select the full 32-layer Llama2 model as our default architecture.

##### *Fine-Tuning Parameters Selection*

In this section, we conduct ablation experiments to identify the key parameters for fine-tuning. Because the embedding layer and output layers are randomly initialized for adaptation to a new domain, they require training. As a baseline, we freeze the entire transformer blocks of pre-trained Llama2 model, treating it as our default architecture for comparison. We then investigate the effects of freezing only the layer normalization and positional embeddings, as well as incorporating fully connected (FC) layers into the set of trainable parameters. The results, shown in Table S5, indicate that adding extra FC layers provides notable benefits for influenza forecasting. With FC layers included, all correlation metrics and percentage-based error metrics (MAPE and SMAPE) consistently improve, whereas absolute-orror metrics such as MAE and MSE exhibit slight increases. Moreover, performance continues to rise as the dimensionality of these FC layers grows. For instance, with an FC-512 layer, SpearmanR and PearsonR improve by 2.81% and 1.06%, respectively, while MAPE and SMAPE decrease by 23.76% and 10.93%. These findings underscore the advantages of larger fully connected layers in boosting correlation and percentage-based error metrics for influenza forecasting.

The results also indicate that re-training the layer normalization and position embeddings (Partial Freeze) provides minimal benefit. Specifically, SpearmanR and PearsonR decrease slightly by 0.24% and 0.12%, respectively, while MAE and MSE remain nearly unchanged compared to the fully frozen Llama2 model. This limited impact may be attributed to the relatively small sample size.

Under our default model framework, we use MSE as the loss function, FP16 for model weight precision, and a learning rate of $1 \times 10^{-4}$. We then explore how alternative design choicessuch as switching to Smooth L1 Loss or Weighted MSE-MAE Loss, changing model weight precision to 8-bit, and introducing batch normalization or data standardizationaffect performance.

The results in Table S6 and Table S7 show that, aside from batch normalization, none of the tested configurations yielded significant differences. Consequently, we maintain MSE as the default loss function, adopt a learning rate of $1 \times 10^{-4}$, and exclude batch normalization and standardization in our default setup. Moreover, Table S6 indicates that reducing model weight precision to 8-bit does not meaningfully affect performance. In resource-constrained scenarios, therefore, 8-bit precision offers a practical alternative, substantially reducing GPU usage while preserving similar model quality.

##### *Detailed Result of Influenza Forecasting on 13-Week prediction length*

The results presented in Table S1 illustrate the predictions of LLMs and other models for the 13-week window from October 2019 to January 2020. Notably, Llama2 outperforms other models across nearly all evaluation metrics, with the exception of the Pearson correlation coefficient.

##### *Detailed Results of Model Performance on Varied prediction lengths*

As shown in Table S8, Table S9 and Table S10, GPT2 and Llama2 exhibit better performance on error metrics but show slightly weaker capabilities in capturing correlation coefficients. The results in Table S8 also show that the performance of LLMs improves steadily as the prediction length increases, while classical models such as SARIMA may experience a decline, as observed in the ILI datasets for Southern and Northern China.

#### A.3 Supplementary Tables

**Table S1:** Comparative analysis of forecasting models for influenza positivity rate prediction in Chongqing. The evaluation is conducted over a single 13-week period from October 2019 to January 2020, ensuring a consistent temporal scope without averaging across multiple runs. **Black**: best. The corresponding model predictions are visualized in Figure 2.

| Model | Spearman's $\rho$ | Pearson's $\rho$ | MAE | MSE | MAPE | SMAPE |
|---|---|---|---|---|---|---|
| Fine-tuned Llama2 | **0.918** | 0.851 | **0.087** | **0.015** | **17.033** | **18.577** |
| Fine-tuned GPT2 | 0.896 | **0.862** | 0.121 | 0.026 | 32.278 | 36.293 |
| SARIMA | 0.868 | 0.791 | 0.104 | 0.025 | 21.178 | 22.888 |
| LSTM | 0.896 | 0.809 | 0.102 | 0.025 | 22.641 | 25.357 |
| PatchTST | 0.874 | 0.849 | 0.089 | 0.015 | 17.910 | 19.707 |

**Table S2:** Detailed results on Spearman and Pearson correlation coefficients. The SARIMA model is employed solely for predicting the final 13-week period from October 2019 to January 2020. To account for computational constraints, model weight precision settings are specified as follows: INT8 for fine-tuned Llama3 and fine-tuned Gemma2, FP16 for fine-tuned Llama2, and FP32 for fine-tuned GPT2. **Black**: best, **Red**: second best. Abbreviations: Spe.$\rho$ (Spearman's $\rho$), Pea.$r$ (Pearson's $r$)

| | Models | SARIMA | | LSTM | | PatchTST | | Fine-tuned Llama2 | | Fine-tuned Llama3 | | Fine-tuned GPT2 | | Fine-tuned GPT4-o mini | | Fine-tuned Gemma2 | |
|---|---|---|---|---|---|---|---|---|---|---|---|---|---|---|---|---|---|
| | Metric | Spe.$\rho$ | Pea.$r$ | Spe.$\rho$ | Pea.$r$ | Spe.$\rho$ | Pea.$r$ | Spe.$\rho$ | Pea.$r$ | Spe.$\rho$ | Pea.$r$ | Spe.$\rho$ | Pea.$r$ | Spe.$\rho$ | Pea.$r$ | Spe.$\rho$ | Pea.$r$ |
| Pos.Rate | Southern | **0.863** | **0.982** | 0.801 | 0.857 | 0.738 | 0.793 | **0.850** | **0.880** | 0.764 | 0.808 | 0.769 | 0.836 | 0.550 | 0.560 | 0.815 | 0.858 |
| | Northern | **0.901** | **0.979** | 0.816 | 0.844 | 0.756 | 0.781 | **0.819** | **0.852** | 0.712 | 0.755 | 0.817 | 0.839 | 0.485 | 0.499 | 0.548 | 0.574 |
| | Chongqing | **0.868** | **0.791** | **0.430** | 0.410 | 0.375 | 0.414 | 0.406 | **0.432** | 0.383 | 0.390 | 0.413 | 0.414 | 0.358 | 0.384 | 0.234 | 0.244 |
| | USA | **0.984** | **0.980** | 0.754 | 0.835 | 0.736 | 0.828 | **0.757** | **0.843** | 0.706 | 0.791 | 0.731 | 0.808 | 0.351 | 0.397 | 0.564 | 0.622 |
| ILI | Southern | **0.874** | **0.947** | **0.830** | **0.847** | 0.762 | 0.805 | 0.802 | 0.833 | 0.703 | 0.738 | 0.829 | 0.837 | 0.323 | 0.337 | 0.461 | 0.489 |
| | Northern | 0.830 | **0.947** | **0.880** | **0.902** | 0.797 | 0.835 | **0.874** | 0.888 | 0.810 | 0.833 | 0.845 | 0.876 | 0.485 | 0.499 | 0.565 | 0.588 |
| | Chongqing | **0.714** | **0.795** | 0.419 | 0.361 | 0.463 | 0.477 | 0.352 | 0.324 | 0.331 | 0.309 | **0.686** | **0.717** | 0.074 | 0.048 | 0.204 | 0.203 |

**Table S3:** Detailed Results on MAE and MSE metrics. The SARIMA model is employed solely for predicting the final 13-week period from October 2019 to January 2020. To account for computational constraints, model weight precision settings are specified as follows: INT8 for fine-tuned Llama3 and fine-tuned Gemma2, FP16 for fine-tuned Llama2, and FP32 for fine-tuned GPT2. **Black**: best, **Red**: second best.

| | Models | SARIMA | | LSTM | | PatchTST | | Fine-tuned Llama2 | | Fine-tuned Llama3 | | Fine-tuned GPT2 | | Fine-tuned GPT4-o mini | | Fine-tuned Gemma2 | |
|---|---|---|---|---|---|---|---|---|---|---|---|---|---|---|---|---|---|
| | **Metric** | **MAE** | **MSE** | **MAE** | **MSE** | **MAE** | **MSE** | **MAE** | **MSE** | **MAE** | **MSE** | **MAE** | **MSE** | **MAE** | **MSE** | **MAE** | **MSE** |
| Pos.Rate | Southern | 4.045 | 25.65 | 2.082 | 9.582 | 2.550 | 11.67 | 2.194 | 10.07 | 2.757 | 13.54 | **2.035** | **8.404** | 9.200 | 192.0 | 2.348 | 10.68 |
| | Northern | 4.339 | 33.20 | 2.666 | 14.76 | 2.489 | 11.10 | **2.241** | **10.24** | 2.729 | 13.25 | 2.454 | 13.24 | 9.946 | 209.6 | 6.727 | 72.23 |
| | Chongqing | 0.104 | 0.025 | 0.097 | 0.017 | 0.093 | 0.014 | **0.085** | **0.013** | 0.100 | 0.017 | 0.095 | 0.016 | 0.116 | 0.034 | 0.181 | 0.053 |
| | USA | 2.871 | 14.53 | 1.289 | 2.964 | 1.192 | **2.609** | **1.185** | 2.737 | 1.456 | 3.760 | 1.291 | 3.071 | 5.509 | 227.5 | 3.419 | 18.572 |
| ILI | Southern | 17887 | $6.14 \times 10^8$ | 6595 | $1.34 \times 10^8$ | 6109 | $9.60 \times 10^7$ | 6577 | $1.27 \times 10^8$ | 7913 | $1.47 \times 10^8$ | **5765** | $\mathbf{9.45 \times 10^7}$ | 19138 | $7.25 \times 10^8$ | 16964 | $4.72 \times 10^8$ |
| | Northern | 7354 | $1.03 \times 10^8$ | 3870 | $3.52 \times 10^7$ | 3600 | $\mathbf{2.58 \times 10^7}$ | 3524 | $2.93 \times 10^7$ | 3988 | $3.07 \times 10^7$ | **3476** | $2.68 \times 10^7$ | 9900 | $1.60 \times 10^8$ | 9900 | $1.60 \times 10^8$ |
| | Chongqing | 57.41 | 5703 | **44.37** | **4746** | 73.68 | 8981 | 54.42 | 5803 | 58.38 | 6503 | 72.78 | 9069 | 63.23 | 9746 | 86.80 | 13420 |

**Table S4:** Detailed Results on MAPE and SMAPE metrics. The SARIMA model is employed solely for predicting the final 13-week period from October 2019 to January 2020. To account for computational constraints, model weight precision settings are specified as follows: INT8 for fine-tuned Llama3 and fine-tuned Gemma2, FP16 for fine-tuned Llama2, and FP32 for fine-tuned GPT2. **Black**: best, **Red**: second best.

| | Models | SARIMA | | LSTM | | PatchTST | | Fine-tuned Llama2 | | Fine-tuned Llama3 | | Fine-tuned GPT2 | | Fine-tuned GPT4-o mini | | Fine-tuned Gemma2 | |
|---|---|---|---|---|---|---|---|---|---|---|---|---|---|---|---|---|---|
| | **Metric** | **MAPE** | **SMAPE** | **MAPE** | **SMAPE** | **MAPE** | **SMAPE** | **MAPE** | **SMAPE** | **MAPE** | **SMAPE** | **MAPE** | **SMAPE** | **MAPE** | **SMAPE** | **MAPE** | **SMAPE** |
| Pos.Rate | Southern | 144.2 | 98.59 | **28.35** | **24.14** | 36.86 | 30.65 | 29.63 | 26.68 | 41.04 | 33.17 | 31.35 | 26.66 | 203.6 | 70.79 | 33.90 | 29.00 |
| | Northern | 64.63 | 41.88 | 49.11 | 34.92 | 75.87 | 41.62 | 43.86 | 32.81 | 72.56 | 46.74 | **36.82** | **32.37** | 257.8 | 82.50 | 253.57 | 85.08 |
| | Chongqing | **21.18** | **22.89** | $2.49 \times 10^8$ | 96.17 | $1.87 \times 10^8$ | 102.6 | $1.73 \times 10^8$ | 96.03 | $2.15 \times 10^8$ | 104.9 | $2.45 \times 10^8$ | 98.46 | $8.98 \times 10^7$ | 116.6 | $4.89 \times 10^8$ | 121.68 |
| | USA | 35.39 | 20.84 | 15.34 | 15.57 | 17.10 | 17.63 | **13.96** | **14.04** | 19.89 | 21.67 | 16.36 | 16.75 | 58.70 | 49.99 | 63.89 | 59.62 |
| ILI | Southern | 25.68 | 22.32 | 7.117 | 7.004 | 7.143 | 7.153 | 7.525 | 7.402 | 9.383 | 9.272 | **6.366** | **6.396** | 24.65 | 23.54 | 21.68 | 22.62 |
| | Northern | 16.54 | 15.03 | 7.766 | 7.696 | 8.076 | 8.037 | **7.283** | **7.234** | 8.950 | 8.941 | 7.340 | 7.412 | 25.98 | 27.19 | 25.98 | 27.19 |
| | Chongqing | **19.07** | **19.21** | 24.45 | 23.15 | 26.56 | 22.77 | 33.29 | 32.26 | 35.67 | 36.05 | 25.03 | 22.22 | 39.06 | 35.10 | 56.70 | 58.73 |

**Table S5:** Ablation study on parameter freezing strategies and prediction block architectures. "Partial Freeze" refers to freezing only the layer normalization and positional embeddings in the fine-tuned Llama2. "FC-$X$" denotes a fully connected layer with $X$ dimensions in the prediction block (e.g., FC-128, FC-256, and FC-512). This study is conducted on the positive rate dataset from Northern China to systematically assess the effects of different architectural modifications on model performance. **Black**: best.

| Model | Spearman's $\rho$ | Pearson's $r$ | MAE | MSE | MAPE | SMAPE |
|---|---|---|---|---|---|---|
| Fine-tuned Llama2 | 0.819 | 0.852 | **2.241** | 10.24 | 43.86 | 32.81 |
| Partial Freeze | 0.817 | 0.851 | 2.244 | **10.21** | 44.72 | 33.36 |
| + FC-128 | 0.825 | 0.847 | 2.619 | 15.08 | 35.75 | 31.34 |
| + FC-256 | 0.840 | 0.858 | 2.584 | 14.84 | 34.30 | 29.99 |
| + FC-512 | **0.842** | **0.861** | 2.576 | 14.83 | **33.43** | **29.22** |

**Table S6:** Ablation study on loss functions and data processing techniques. "Smooth L1" combines the advantages of L1 and L2 losses, ensuring stability for small errors while being less sensitive to outliers. "Weighted MSE-MAE" optimally balances MSE and MAE to enhance performance across both small and large errors. "INT8" represents 8-bit model weight quantization, "BN" denotes batch normalization, and "Scale" refers to standardization. This study is conducted on the positive rate dataset from Northern China to systematically evaluate the impact of these modifications on model performance. **Black**: best.

| Model | Spearman's $\rho$ | Pearson's $r$ | MAE | MSE | MAPE | SMAPE |
|---|---|---|---|---|---|---|
| MSE | 0.819 | 0.852 | **2.241** | **10.24** | 43.86 | 32.81 |
| Smooth L1 | 0.819 | 0.847 | 2.261 | 10.65 | 42.70 | **32.62** |
| Weighted MSE-MAE | 0.823 | 0.853 | 2.243 | 10.25 | 44.02 | 33.04 |
| INT8 | 0.809 | 0.843 | 2.310 | 10.80 | 44.31 | 33.17 |
| + BN | 0.792 | 0.823 | 2.339 | 11.10 | **42.29** | 39.76 |
| + Scale | **0.825** | **0.859** | 2.293 | 10.68 | 44.12 | 34.10 |

**Table S7:** Ablation study on the learning rate configuration of the fine-tuned Llama2 model. This study systematically examines the influence of varying learning rates on model performance using the positive rate dataset from Northern China. **Black**: best.

| Metric | Spearman's $\rho$ | Pearson's $r$ | MAE | MSE | MAPE | SMAPE |
|---|---|---|---|---|---|---|
| **LR $1 \times 10^{-4}$** | **0.819** | **0.852** | **2.241** | **10.24** | **43.86** | **32.81** |
| **LR $1 \times 10^{-3}$** | 0.790 | 0.824 | 2.312 | 10.59 | 51.56 | 37.70 |
| **LR $1 \times 10^{-5}$** | 0.775 | 0.800 | 2.955 | 17.59 | 56.72 | 35.83 |

**Table S8:** Comparison of models based on MAE and MSE metrics. Results are presented for prediction lengths of 8 and 13. **Black**: best, **Red**: second best.

| | Models | | SARIMA | | LSTM | | PatchTST | | Fine-tuned Llama2 | | Fine-tuned GPT2 | |
|---|---|---|---|---|---|---|---|---|---|---|---|---|
| | Metric | | MAE | MSE | MAE | MSE | MAE | MSE | MAE | MSE | MAE | MSE |
| Pos.Rate | Southern | 8 | 4.068 | 21.94 | **2.094** | **9.823** | 2.592 | 12.53 | 2.369 | 11.41 | 2.241 | 11.14 |
| | | 13 | 4.045 | 25.65 | 2.082 | 9.582 | 2.550 | 11.67 | 2.194 | 10.07 | **2.035** | **8.404** |
| | Northern | 8 | 6.736 | 48.71 | 2.709 | 14.97 | 2.645 | 12.14 | **2.293** | **10.70** | 2.655 | 15.84 |
| | | 13 | 4.339 | 33.20 | 2.666 | 14.76 | 2.489 | 11.10 | **2.241** | **10.24** | 2.454 | 13.24 |
| | Chongqing | 8 | 0.100 | 0.024 | 0.095 | 0.016 | 0.089 | 0.014 | **0.084** | **0.013** | 0.108 | 0.020 |
| | | 13 | 0.104 | 0.025 | 0.097 | 0.017 | 0.093 | 0.014 | **0.085** | **0.013** | 0.095 | 0.016 |
| | USA | 8 | 3.150 | 16.42 | 1.268 | 2.840 | **1.127** | **2.460** | 1.194 | 2.831 | 1.376 | 3.610 |
| | | 13 | 2.871 | 14.53 | 1.289 | 2.964 | 1.192 | **2.609** | **1.185** | 2.737 | 1.291 | 3.071 |
| ILI | Southern | 8 | 12500 | 1.86e8 | **6720** | **1.40e8** | 7373 | $1.43 \times 10^8$ | 7007 | $1.44 \times 10^8$ | 6844 | $1.43 \times 10^8$ |
| | | 13 | 17887 | $6.14 \times 10^8$ | 6595 | $1.34 \times 10^8$ | 6109 | $9.60 \times 10^7$ | 6577 | $1.27 \times 10^8$ | **5765** | **$9.45 \times 10^7$** |
| | Northern | 8 | **3180** | **$1.39 \times 10^7$** | 3890 | $3.58 \times 10^7$ | 3766 | $3.22 \times 10^7$ | 3806 | $3.45 \times 10^7$ | 3961 | $3.71 \times 10^7$ |
| | | 13 | 7354 | $1.03 \times 10^8$ | 3870 | $3.52 \times 10^7$ | 3600 | **$2.58 \times 10^7$** | 3524 | $2.93 \times 10^7$ | **3476** | $2.68 \times 10^7$ |
| | Chongqing | 8 | 70.03 | 8633 | **44.74** | **4716** | 51.98 | 5354 | 49.83 | 5036 | 45.23 | 4740 |
| | | 13 | 57.41 | 5703 | **44.37** | **4746** | 73.68 | 8981 | 54.42 | 5803 | 72.78 | 9069 |

**Table S9:** Comparison of models bsed on MAPE and SMAPE metrics. Results are presented for prediction lengths of 8 and 13. **Black**: best, **Red**: second best.

| | Models | | SARIMA | | LSTM | | PatchTST | | Fine-tuned Llama2 | | Fine-tuned GPT2 | |
|---|---|---|---|---|---|---|---|---|---|---|---|---|
| | Metric | | MAPE | SMAPE | MAPE | SMAPE | MAPE | SMAPE | MAPE | SMAPE | MAPE | SMAPE |
| Pos.Rate | Southern | 8 | 292.2 | 176.3 | **32.80** | **26.45** | 41.06 | 34.75 | 39.50 | 34.19 | 38.18 | 32.36 |
| | | 13 | 144.2 | 98.59 | **28.35** | **24.14** | 36.86 | 30.65 | 29.63 | 26.68 | 31.35 | 26.66 |
| | Northern | 8 | 256.2 | 183.1 | 58.00 | 38.71 | 77.79 | 43.96 | 47.55 | 37.01 | **37.09** | **32.23** |
| | | 13 | 64.63 | 41.88 | 49.11 | 34.92 | 75.87 | 41.62 | 43.86 | 32.81 | **36.82** | **32.37** |
| | Chongqing | 8 | **17.99** | **16.15** | 2.45e8 | 95.91 | 1.79e8 | 100.3 | 1.65e8 | 93.56 | 2.88e8 | 104.0 |
| | | 13 | **21.18** | **22.89** | 2.49e8 | 96.17 | 1.87e8 | 102.6 | 1.73e8 | 96.03 | 2.45e8 | 98.46 |
| | USA | 8 | 47.38 | 27.94 | 15.97 | 16.17 | 14.72 | 15.01 | **14.09** | 14.19 | 14.63 | **14.10** |
| | | 13 | 35.39 | 20.84 | 15.34 | 15.57 | 17.10 | 17.63 | **13.96** | **14.04** | 16.36 | 16.75 |
| ILI | Southern | 8 | 43.15 | 58.19 | **7.468** | **7.303** | 8.899 | 8.713 | 8.304 | 8.058 | 7.646 | 7.482 |
| | | 13 | 25.68 | 22.32 | 7.117 | 7.004 | 7.143 | 7.153 | 7.525 | 7.402 | **6.366** | **6.396** |
| | Northern | 8 | 14.54 | 13.80 | **8.006** | **7.900** | 8.093 | 7.966 | 8.107 | 7.953 | 8.195 | 8.084 |
| | | 13 | 16.54 | 15.03 | 7.766 | 7.696 | 8.076 | 8.037 | **7.283** | **7.234** | 7.340 | 7.412 |
| | Chongqing | 8 | **21.15** | **20.52** | 40.15 | 28.44 | 29.88 | 29.30 | 28.61 | 27.46 | 24.72 | 23.63 |
| | | 13 | **19.07** | **19.21** | 24.45 | 23.15 | 26.56 | 22.77 | 33.29 | 32.26 | 25.03 | 22.22 |

**Table S10:** Comparison of models based on correlation coefficients. Results are presented for prediction lengths of 8 and 13. **Note:** Spe.$\rho$ and Pea.$r$ refer to Spearman's $\rho$ and Pearson's $r$, respectively. **Black**: best, **Red**: second best.

| | Models | | SARIMA | | LSTM | | PatchTST | | Fine-tuned Llama2 | | Fine-tuned GPT2 | |
|---|---|---|---|---|---|---|---|---|---|---|---|---|
| | Metric | | Spe.$\rho$ | Pea.$r$ | Spe.$\rho$ | Pea.$r$ | Spe.$\rho$ | Pea.$r$ | Spe.$\rho$ | Pea.$r$ | Spe.$\rho$ | Pea.$r$ |
| Pos.Rate | Southern | 8 | 0.405 | **0.914** | 0.644 | 0.706 | 0.665 | 0.713 | **0.711** | 0.769 | 0.689 | 0.747 |
| | | 13 | **0.863** | **0.982** | 0.801 | 0.857 | 0.738 | 0.793 | 0.850 | 0.880 | 0.769 | 0.836 |
| | Northern | 8 | 0.595 | **0.848** | 0.772 | 0.776 | 0.706 | 0.714 | 0.761 | 0.771 | **0.799** | 0.805 |
| | | 13 | **0.901** | **0.979** | 0.816 | 0.844 | 0.756 | 0.781 | 0.819 | 0.852 | 0.817 | 0.839 |
| | Chongqing | 8 | **0.667** | **0.546** | 0.336 | 0.248 | 0.345 | 0.314 | 0.343 | 0.296 | 0.291 | 0.229 |
| | | 13 | **0.868** | **0.791** | 0.430 | 0.410 | 0.375 | 0.414 | 0.406 | 0.432 | 0.413 | 0.414 |
| | USA | 8 | **1.000** | **0.981** | 0.642 | 0.724 | 0.642 | 0.732 | 0.629 | 0.724 | 0.654 | 0.731 |
| | | 13 | **0.984** | **0.980** | 0.754 | 0.835 | 0.736 | 0.828 | 0.757 | 0.843 | 0.731 | 0.808 |
| ILI | Southern | 8 | 0.286 | **0.888** | 0.717 | 0.757 | 0.645 | 0.692 | 0.688 | 0.717 | **0.721** | 0.755 |
| | | 13 | **0.874** | **0.947** | 0.830 | 0.847 | 0.762 | 0.805 | 0.802 | 0.833 | 0.829 | 0.837 |
| | Northern | 8 | 0.095 | 0.810 | **0.782** | **0.827** | 0.752 | 0.782 | 0.780 | 0.810 | 0.763 | 0.818 |
| | | 13 | 0.830 | **0.947** | **0.880** | 0.902 | 0.797 | 0.835 | 0.874 | 0.888 | 0.845 | 0.876 |
| | Chongqing | 8 | 0.381 | 0.568 | **0.592** | **0.633** | 0.238 | 0.225 | 0.265 | 0.250 | 0.268 | 0.248 |
| | | 13 | **0.714** | **0.795** | 0.419 | 0.361 | 0.463 | 0.477 | 0.352 | 0.324 | 0.686 | 0.717 |

**Table S11:** Zero-shot learning results from three replications. Each model generates predictions without prior training on the related datasets.

| | Models | GPT2 | | | | | | Llama2 | | | | | |
|---|---|---|---|---|---|---|---|---|---|---|---|---|---|
| | Metric | Spe.$\rho$ | Pea.$r$ | MAE | MSE | MAPE | SMAPE | Spe.$\rho$ | Pea.$r$ | MAE | MSE | MAPE | SMAPE |
| Pos.Rate | Southern | 0.848 | 0.980 | 4.281 | 33.07 | 112.22 | 76.79 | 0.868 | 0.981 | 4.086 | 31.03 | 111.8 | 66.51 |
| | Northern | 0.853 | 0.979 | 4.207 | 35.34 | 55.70 | 36.53 | 0.830 | 0.978 | 4.385 | 34.86 | 68.30 | 43.50 |
| | Chongqing | 0.886 | 0.808 | 0.105 | 0.027 | 23.29 | 26.13 | 0.852 | 0.750 | 0.134 | 0.036 | 28.38 | 31.59 |
| | USA | 0.967 | 0.977 | 2.762 | 11.66 | 23.10 | 17.53 | 0.947 | 0.967 | 2.732 | 13.23 | 20.51 | 16.31 |
| ILI | Southern | 0.855 | 0.948 | 15672 | $5.06 \times 10^8$ | 20.84 | 18.62 | 0.863 | 0.948 | 16024 | $4.84 \times 10^8$ | 22.70 | 20.35 |
| | Northern | 0.802 | 0.954 | 8728 | $1.18 \times 10^8$ | 21.07 | 19.04 | 0.819 | 0.951 | 9051 | $1.20 \times 10^8$ | 23.25 | 21.21 |
| | Chongqing | 0.936 | 0.953 | 145.9 | 32112 | 20.57 | 23.51 | 0.703 | 0.764 | 65.45 | 7066 | 21.49 | 22.44 |

**Table S12:** Comparison of pre-trained Llama2 and SARIMA. The values in the table represent the percentage improvement of pre-trained Llama2 over SARIMA. The improvement rate for all metrics is computed as [(pre-trained Llama2 − SARIMA)/SARIMA] $\times$ 100%. To emphasize central tendencies in performance variations, the median improvement value for each metric across different datasets is underlined.

| | Dataset | Spearman's $\rho$ (%) | Pearson's $r$ (%) | MAE (%) | MSE (%) | MAPE (%) | SMAPE (%) |
|---|---|---|---|---|---|---|---|
| Pos.Rate | Southern | 0.58 | <u>-0.10</u> | 1.01 | 20.97 | -22.47 | -32.54 |
| | Northern | -7.88 | -0.10 | <u>1.06</u> | 5.00 | <u>5.68</u> | <u>3.87</u> |
| | Chongqing | -1.84 | -5.18 | 28.85 | 44.00 | 33.99 | 38.01 |
| | USA | -3.76 | -1.33 | -4.84 | -8.95 | -42.05 | -21.74 |
| ILI | Southern | -1.26 | 0.11 | -10.42 | -21.17 | -11.60 | -8.83 |
| | Northern | -1.33 | 0.42 | 23.08 | <u>16.50</u> | 40.57 | 41.12 |
| | Chongqing | <u>-1.54</u> | -3.90 | 14.00 | 23.90 | 12.69 | 16.81 |

**Table S13:** Comparison of pre-trained GPT-2 and SARIMA. The values in the table represent the percentage improvement of pre-trained GPT-2 over SARIMA. The improvement rate for all metrics is computed as [(pre-trained GPT-2 − SARIMA)/SARIMA] $\times$ 100%. To emphasize central tendencies in performance variations, the median improvement value for each metric across different datasets is underlined.

| | Dataset | Spearman's $\rho$ (%) | Pearson's $r$ (%) | MAE (%) | MSE (%) | MAPE (%) | SMAPE (%) |
|---|---|---|---|---|---|---|---|
| Pos.Rate | Southern | <u>-1.74</u> | -0.20 | 5.83 | 28.93 | -22.18 | -22.11 |
| | Northern | -5.33 | 0.00 | -3.04 | 6.45 | <u>-13.82</u> | <u>-12.77</u> |
| | Chongqing | 2.07 | 2.15 | <u>0.96</u> | <u>8.00</u> | 9.96 | 14.15 |
| | USA | -1.73 | -0.31 | -3.80 | -19.75 | -34.73 | -15.88 |
| ILI | Southern | -2.17 | <u>0.11</u> | -12.38 | -17.59 | -18.85 | -16.58 |
| | Northern | -3.37 | 0.74 | 18.68 | 14.56 | 27.39 | 26.68 |
| | Chongqing | 31.09 | 19.87 | 154.14 | 463.07 | 7.87 | 22.38 |

**Table S14:** Comparison of pre-trained Llama2 and LSTM. The values in the table represent the percentage improvement of pre-trained Llama2 over LSTM. The improvement rate for all metrics is computed as $[(\text{pre-trained Llama2} - \text{LSTM})/\text{LSTM}] \times 100\%$. To emphasize central tendencies in performance variations, the median improvement value for each metric across different datasets is underlined.

| | Dataset | Spearman's $\rho$ (%) | Pearson's $r$ (%) | MAE (%) | MSE (%) | MAPE (%) | SMAPE (%) |
|---|---|---|---|---|---|---|---|
| Pos.Rate | Southern | 8.36 | 14.47 | 96.25 | 223.9 | 294.36 | 175.52 |
| | Northern | 1.72 | 15.88 | 64.48 | 136.18 | 39.08 | 24.57 |
| | Chongqing | 98.14 | 82.93 | 38.14 | 111.76 | -100.00 | -67.16 |
| | USA | 25.60 | 15.81 | 111.95 | 346.36 | 33.70 | 4.75 |
| ILI | Southern | 3.98 | 11.92 | 142.97 | 261.19 | 218.82 | 190.71 |
| | Northern | -6.93 | 5.43 | 133.88 | 240.91 | 199.23 | 175.45 |
| | Chongqing | 67.78 | 111.63 | 47.51 | 48.88 | -12.11 | -3.07 |

**Table S15:** Comparison of pre-trained GPT-2 and LSTM. The values in the table represent the percentage improvement of pre-trained GPT-2 over LSTM. The improvement rate for all metrics is computed as $[(\text{pre-trained GPT-2} - \text{LSTM})/\text{LSTM}] \times 100\%$. To emphasize central tendencies in performance variations, the median improvement value for each metric across different datasets is underlined.

| | Dataset | Spearman's $\rho$ (%) | Pearson's $r$ (%) | MAE (%) | MSE (%) | MAPE (%) | SMAPE (%) |
|---|---|---|---|---|---|---|---|
| Pos.Rate | Southern | 5.87 | 14.35 | 105.62 | 245.2 | 295.84 | 218.1 |
| | Northern | 4.53 | 16.00 | 57.80 | 139.43 | 13.42 | 4.61 |
| | Chongqing | 106.05 | 97.07 | 8.25 | 58.82 | -100.00 | -72.84 |
| | USA | 28.25 | 17.01 | 114.27 | 293.39 | 50.59 | 12.59 |
| ILI | Southern | 3.01 | 11.92 | 137.63 | 277.61 | 192.70 | 166.00 |
| | Northern | -8.86 | 5.76 | 125.53 | 235.23 | 171.17 | 147.27 |
| | Chongqing | 123.39 | 163.99 | 228.83 | 576.61 | -15.87 | 1.56 |

**Table S16:** Comparison of pre-trained Llama2 and PatchTST. The values in the table represent the percentage improvement of pre-trained Llama2 over PatchTST. The improvement rate for all metrics is computed as $[(\text{pre-trained Llama2} - \text{PatchTST})/\text{PatchTST}] \times 100\%$. To emphasize central tendencies in performance variations, the median improvement value for each metric across different datasets is underlined.

| | Dataset | Spearman's $\rho$ (%) | Pearson's $r$ (%) | MAE (%) | MSE (%) | MAPE (%) | SMAPE (%) |
|---|---|---|---|---|---|---|---|
| Pos.Rate | Southern | 17.62 | 23.71 | 60.24 | 165.9 | 203.31 | 117.00 |
| | Northern | 9.79 | 25.22 | 76.18 | 214.05 | -9.98 | 4.52 |
| | Chongqing | 127.20 | 81.16 | 44.09 | 157.14 | -100.00 | -69.21 |
| | USA | 28.67 | 16.79 | 129.19 | 407.09 | 19.94 | -7.49 |
| ILI | Southern | 13.25 | 17.76 | 162.30 | 404.17 | 217.79 | 184.50 |
| | Northern | 2.76 | 13.89 | 151.42 | 365.12 | 187.89 | 163.90 |
| | Chongqing | 51.84 | 60.17 | -11.17 | -21.32 | -19.09 | -1.45 |

**Table S17:** Comparison of pre-trained GPT-2 and PatchTST. The values in the table represent the percentage improvement of pre-trained GPT-2 over PatchTST. The improvement rate for all metrics is computed as $[(\text{pre-trained GPT-2} - \text{PatchTST})/\text{PatchTST}] \times 100\%$. To emphasize central tendencies in performance variations, the median improvement value for each metric across different datasets is underlined.

| | Dataset | Spearman's $\rho$ (%) | Pearson's $r$ (%) | MAE (%) | MSE (%) | MAPE (%) | SMAPE (%) |
|---|---|---|---|---|---|---|---|
| Pos.Rate | Southern | 14.91 | 23.58 | 67.88 | 183.38 | 204.45 | 150.54 |
| | Northern | 12.83 | 25.35 | 69.02 | 218.38 | -26.58 | -12.23 |
| | Chongqing | 136.27 | 95.17 | 12.90 | 92.86 | -100.00 | -74.53 |
| | USA | 31.39 | 18.00 | 131.71 | 346.91 | 35.09 | -0.57 |
| ILI | Southern | 12.20 | 17.76 | 156.54 | 427.08 | 191.75 | 160.31 |
| | Northern | 0.63 | 14.25 | 142.44 | 357.36 | 160.90 | 136.90 |
| | Chongqing | 102.16 | 99.79 | 98.02 | 257.55 | -22.55 | 3.25 |

**Table S18:** Comparison of pre-trained Llama2 and fine-tuned Llama2. The values in the table represent the percentage improvement of pre-trained Llama2 over fine-tuned Llama2. The improvement rate for all metrics is computed as $[(\text{pre-trained Llama2} - \text{fine-tuned Llama2})/\text{fine-tuned Llama2}] \times 100\%$. To emphasize central tendencies in performance variations, the median improvement value for each metric across different datasets is underlined.

| Dataset | Spearman's $\rho$ (%) | Pearson's $r$ (%) | MAE (%) | MSE (%) | MAPE (%) | SMAPE (%) |
|---|---|---|---|---|---|---|
| Southern | 2.12 | 11.48 | 86.24 | 208.14 | 277.32 | 149.29 |
| Northern | 1.34 | 14.79 | 95.67 | 240.43 | 55.72 | 32.58 |
| Chongqing | 109.85 | 73.61 | 57.65 | 176.92 | -100.00 | -67.10 |
| USA | 25.10 | 14.71 | 130.55 | 383.38 | 46.92 | 16.17 |
| Southern | 7.61 | 13.81 | 143.64 | 281.10 | 201.66 | 174.93 |
| Northern | -6.29 | 7.09 | 156.84 | 309.56 | 219.24 | 193.20 |
| Chongqing | 99.72 | 135.80 | 20.27 | 21.76 | -35.45 | -30.44 |

**Table S19:** Comparison of pre-trained GPT-2 and fine-tuned GPT-2. The values in the table represent the percentage improvement of pre-trained GPT-2 over fine-tuned GPT-2. The improvement rate for all metrics is computed as $[(\text{pre-trained GPT-2} - \text{fine-tuned GPT-2})/\text{fine-tuned GPT-2}] \times 100\%$. To emphasize central tendencies in performance variations, the median improvement value for each metric across different datasets is underlined.

| Dataset | Spearman's $\rho$ (%) | Pearson's $r$ (%) | MAE (%) | MSE (%) | MAPE (%) | SMAPE (%) |
|---|---|---|---|---|---|---|
| Southern | 10.27 | 17.22 | 110.37 | 293.50 | 257.96 | 188.03 |
| Northern | 4.41 | 16.69 | 71.43 | 166.92 | 51.28 | 12.85 |
| Chongqing | 32.28 | 20.92 | 113.94 | 279.68 | 41.20 | 4.66 |
| USA | 114.53 | 95.17 | 10.53 | 68.75 | -100.00 | -73.46 |
| Southern | 3.14 | 13.26 | 171.85 | 435.45 | 227.36 | 191.12 |
| Northern | -5.09 | 8.90 | 151.09 | 340.30 | 187.06 | 156.88 |
| Chongqing | 36.44 | 32.91 | 100.47 | 254.09 | -17.82 | 5.81 |

413 ## A.4 Supplementary Figures

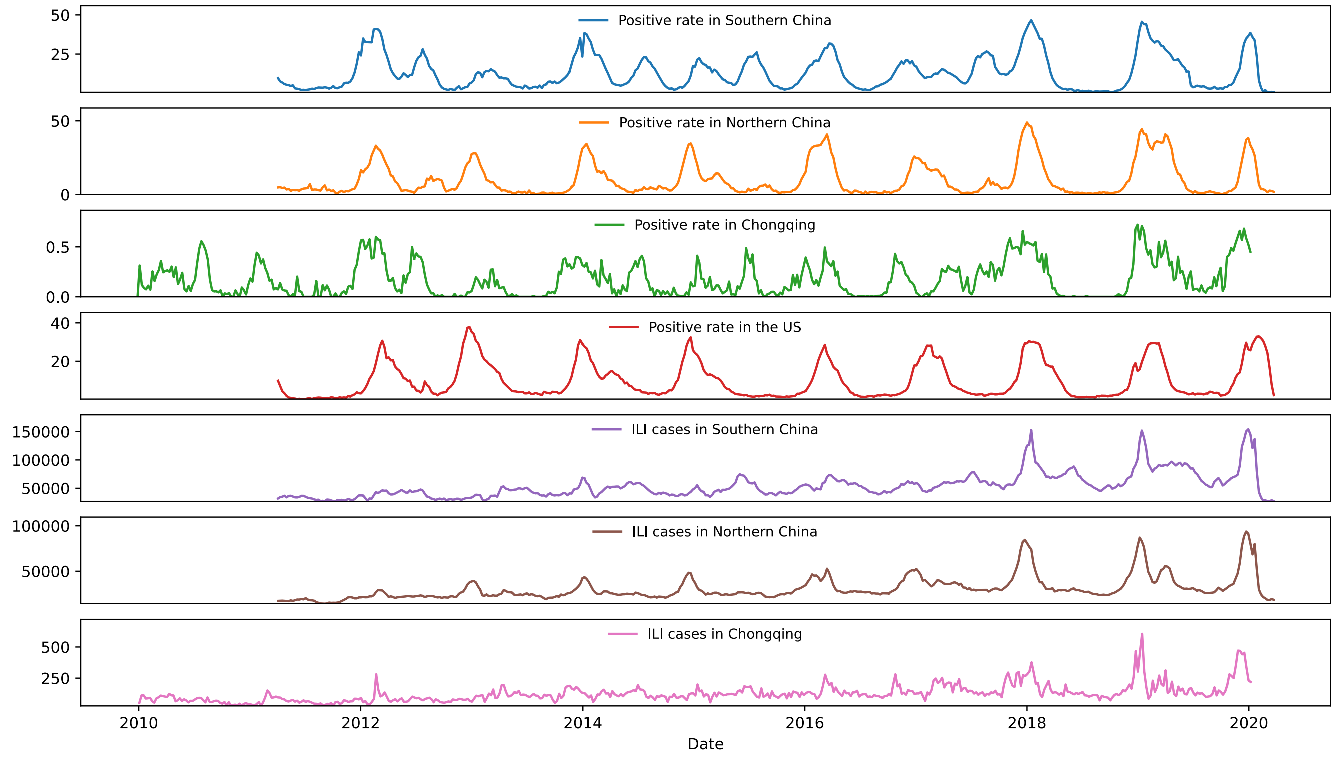


**Figure S1:** Temporal dynamics of influenza activity as captured by the surveillance datasets analyzed in this study. The upper four panels display the weekly positive detection rates of influenza viruses in southern China, northern China, Chongqing, and the United States, respectively. The lower four panels present the corresponding weekly counts of ILI cases. Data for southern China, northern China, and the United States are obtained from [13].

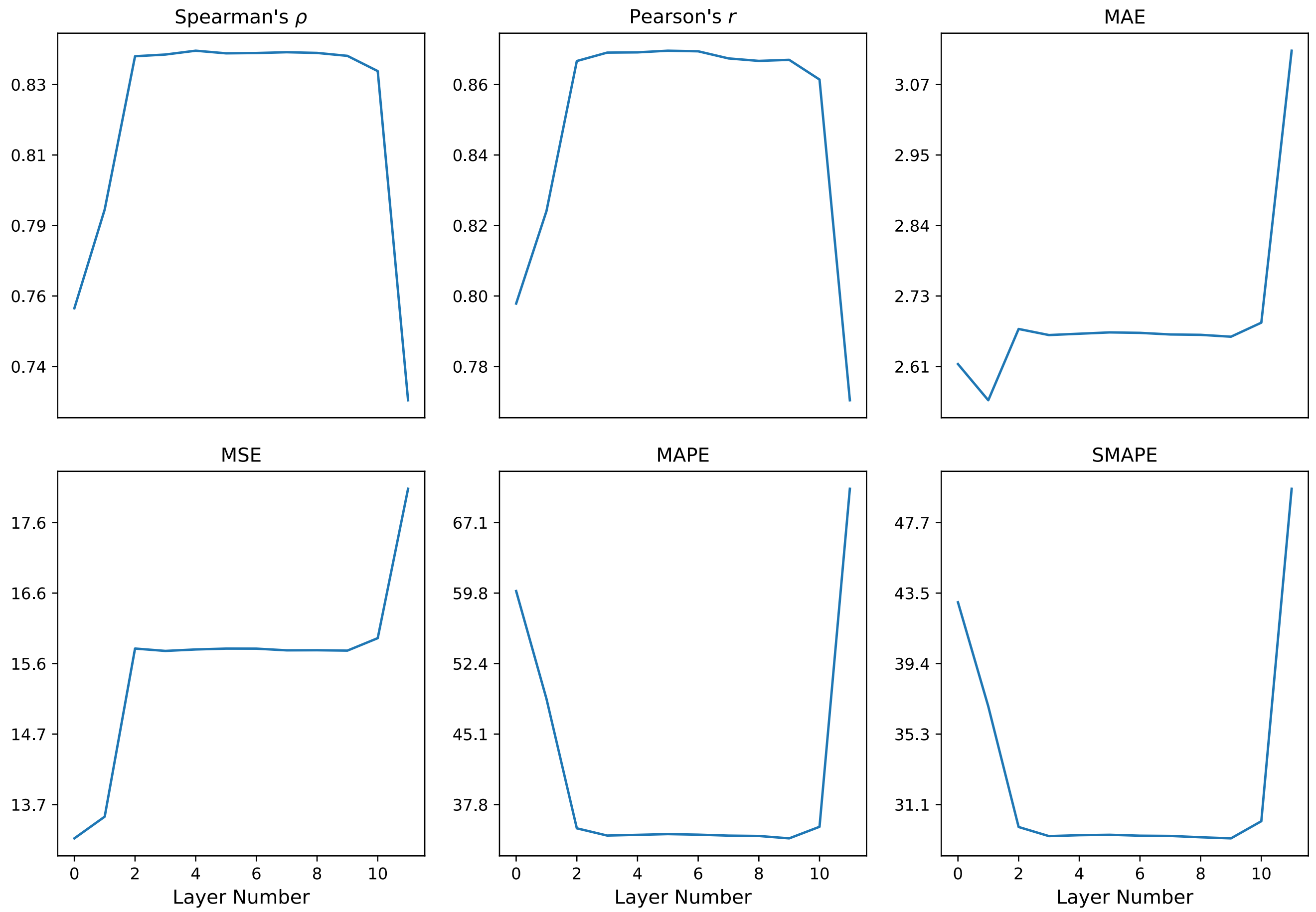


**Figure S2:** Comparison of pre-trained GPT2 model with different numbers of layers on the positive rate dataset from Northern China across all metrics.

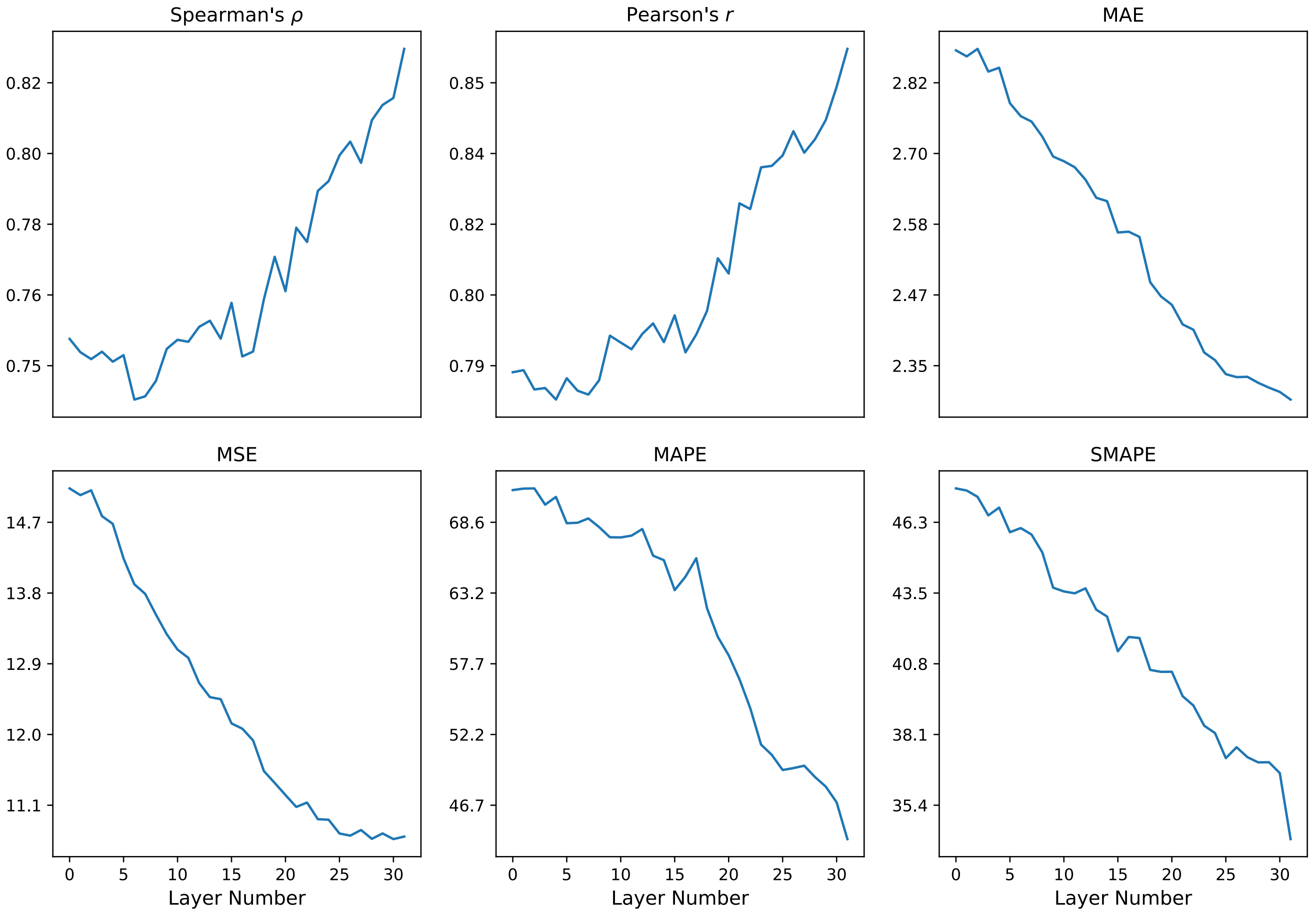


**Figure S3:** Comparison of pre-trained Llama2 model with different numbers of layers on the positive rate dataset from Northern China across all metrics.
